# Disease association with frequented regions of genotype graphs

**DOI:** 10.1101/2020.09.25.20201640

**Authors:** Samuel Hokin, Alan Cleary, Joann Mudge

**Affiliations:** National Center for Genome Resources, Santa Fe, New Mexico, USA

## Abstract

Complex diseases, with many associated genetic and environmental factors, are a challenging target for genomic risk assessment. Genome-wide association studies (GWAS) associate disease status with, and compute risk from, individual common variants, which can be problematic for diseases with many interacting or rare variants. In addition, GWAS typically employ a reference genome which is not built from the subjects of the study, whose genetic background may differ from the reference and whose genetic characterization may be limited. We present a complementary method based on disease association with collections of genotypes, called frequented regions, on a pangenomic graph built from subjects’ genomes. We introduce the pangenomic genotype graph, which is better suited than sequence graphs to human disease studies. Our method draws out collections of features, across multiple genomic segments, which are associated with disease status. We show that the frequented regions method consistently improves machine-learning classification of disease status over GWAS classification, allowing incorporation of rare or interacting variants. Notably, genomic segments that have few or no variants of genome-wide significance (*p* < 5 × 10^−8^) provide much-improved classification with frequented regions, encouraging their application across the entire genome. Frequented regions may also be utilized for purposes such as choice of treatment in addition to prediction of disease risk.

## INTRODUCTION

Complex diseases, with many associated genetic and environmental factors, present a challenging diagnostic landscape.[1,2] It is difficult to assess disease risk from genomic data due to the polygenic nature of associations.[3] Genome-wide association studies (GWAS) are standard measures of genomic association with disease[4], but they typically treat associated features, often single-nucleotide polymorphisms (SNPs), independently, and assess only common variants even though rare alleles collectively represent a much larger pool of disease risk loci.[5] A whole-genome approach is motivated by the fact that GWAS often find strong associations with variants on genes and intergenic segments on many chromosomes, as well as the observation that these features often appear to interact to enhance disease risk.[3,6–8] A method which emphasizes the combined effect of disease-associated genomic features across the entire genome would provide a complementary and illuminating approach.

Though methods exist to analyze combinations of features from GWAS, the number of variant interaction combinations leads to computational limitations that often require filtering out variant combinations up front based on biological or statistical criteria.[9,10] While computational complexity increases when considering interactions among rare as well as common variants, including rare variants and their interactions can increase predictive power and identify clinically-relevant variants for common diseases.[10,11] Moving from a pairwise variant analysis to analyses based on genomic segments and application of machine learning techniques have both been proposed to overcome computational challenges in identifying interactions between variants or genes and for improving disease prediction.[12–15] Here, we apply a new region-based method and machine learning to link common and rare variants to human disease.

A promising type of genomic feature for this purpose is termed a *frequented region*, which groups genomic segments shared, or partly shared, among sample subsets. First introduced by Cleary *et al*.[16], frequented regions (FRs) are regions of a pangenomic graph that frequently occur within a subset of the population. Here we demonstrate the application of FRs to case-control genomic studies of human disease.

Pangenome graphs[17] are powerful ways to represent genomic variation within a cohort of individuals without resorting to variant calls against a reference genome or genotyping chips that cover only a fraction of the variation in a population. A graph may be built from individuals’ assembled genomes or even directly from whole-genome sequencing (WGS) reads. This is currently an area of active research.[18,19]

A *pangenome-wide association study* (PWAS) using FRs has been presented for 49 traits across 100 strains of yeast[20]. That study employed machine learning of FRs to predict the traits associated with each of the 100 strain genomes. In this report, we apply FRs to human case-control disease studies, where we have many similar but distinct genomes, labeled “case” or “control” depending on disease status of the individual. We also apply machine learning of FRs, in this case to predict disease status.

While FRs provide insight into polygenic associations with disease, our goal is to develop an assay which can estimate the risk of disease in undiagnosed individuals as a complement to conventional polygenic risk scores[6]. Another application would be to guide treatment of affected individuals, by predicting the success of medications when multiple choices are available. Frequented regions provide a natural framework for machine learning (ML)[16,20], in particular for supervised classification[21], so we present results of supervised classification using the support vector machine (SVM) method in our example of a complex disease, schizophrenia.

We present three example case-control analyses to illustrate the methodology and demonstrate its potential benefits: of sickle-cell disease (SCD), Huntington disease (HD), and schizophrenia (SCZ). The first two examples highlight distinct aspects of the method on Mendelian diseases for explanatory purposes; the third example, of a highly complex heritable disease, demonstrates that supervised classification using frequented regions performs better than classification using the same variants treated independently (GWAS).

## METHODS

### Frequented regions and application to case-control disease studies

Frequented regions linked to disease are collections of genomic features which highlight multi-genotypic or polygenic associations with a disease; a frequented region is a portion of a pangenome graph that represents sequence that is approximately conserved in the genomes that support that region. One *searches* for FRs of a pangenomic graph, guided by parameters chosen to bring out a desired type of collection: adjacent variants, multi-allelic variants, or non-adjacent variants on distant segments. The conceptual basis of FRs is that complex diseases are associated with a variety of features across the entire genome, while any given individual’s genome contains a subset of those associated features. By enumerating the FRs on a pangenomic graph, one collects relevant combinations of features for all of the individuals. While a GWAS uses a single variant as an input to risk prediction, an FR-based prediction uses a single FR, composed of many variants, as an input. As there can be thousands of individual variants associated with a disease, there can be thousands of FRs associated with a disease. The particular combination of variants will vary based on the individual genome, and many of the control-labeled genomes will also share some of the causative loci. Therefore, it is important to consider large sets of genomes. Our application of FRs is therefore a method in pangenome-wide association studies (PWAS).

In the original formulation[16], the pangenome graph is built from genomic sequences, and the graph is termed a *sequence graph*. Each vertex, or *node* on the graph represents a genomic segment, perhaps many bases long, that is carried by one or more individuals. The frequented region is designated by a *cluster* of nodes, and the task at hand is to find FRs that are deemed *interesting*, i.e. which satisfy some desirable characteristic, such as consensus among individuals or, in the application reported here, association with affected versus unaffected individuals in a case-control disease study. Two parameters, *α* and *κ*, described below, further specify FRs.

Each individual’s genome presents a *path* or paths through the graph. On a sequence graph, each diploid individual has two paths, which require phased assembly[22]. We introduce an alternative pangenome *genotype graph*, on which nodes represent genotypes, perhaps many bases long, rather than sequence (alleles) and an individual is represented by a single path through the graph. This typically increases the number of nodes (the genotypes `A/A`, `A/T`, and `T/T` are three nodes on a genotype graph, while occupying two nodes, `A` and `T`, on a sequence graph), while halving the number of paths. Importantly, genotype is associated with disease, and we are interested in the analysis of disease. ([23] presents a statistical argument for using genotypes rather than alleles in disease association studies, as the latter presumes Hardy-Weinberg equilibrium in the controls, which is often not the case.) Figure 1 compares a sequence graph with a genotype graph for the first 400 bases of the *HTT* gene from 37 individuals, 27 cases and 10 controls, in a study of Huntington disease (dbGaP study accession phs001071.v1.p1), both built from the provided variant calls against the GRCh37 human genome reference sequence.

**Figure 1:**
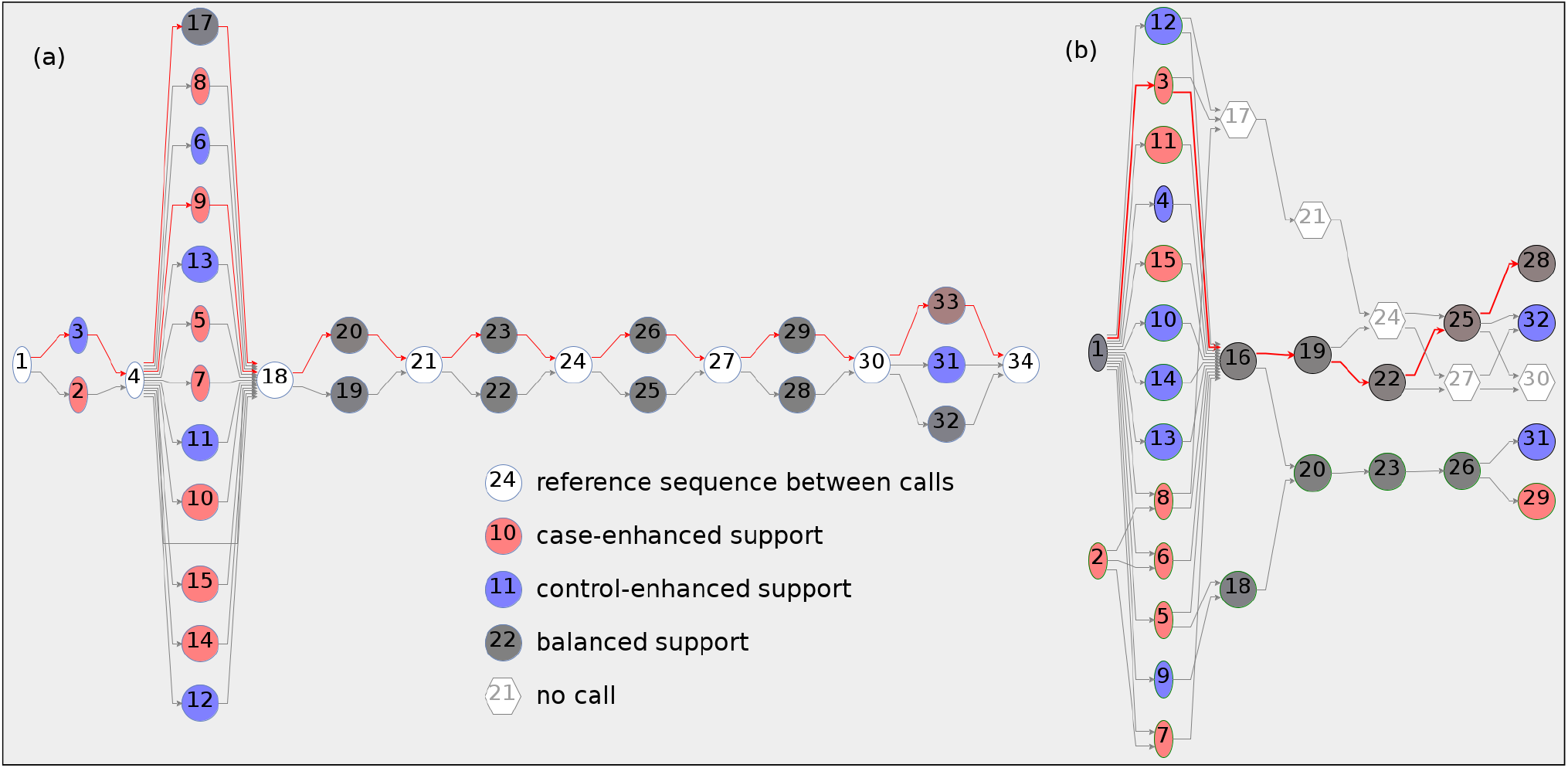
Pangenome graphs of the first 400 bases of the HTT gene from variant calls on 27 case and 10 control subjects in a study of Huntington disease (HD) (dbGaP Study Accession phs001071.v1.p1): **(a)** sequence graph, in which each (diploid) individual has two paths; **(b)** genotype graph, in which each individual has one path. Nodes are numbered consecutively as the graphs are built for identification in analysis; nodes at the same locus are shown vertically. The red line displays the path(s) of an HD-affected individual. In (a), that individual’s paths traverse nodes 9 (`CAG`×41) and 17 (`CAG`×19), with node 9 representing a disease-associated `CAG`-repeat allele. In (b), that individual’s path traverses node 3, corresponding to the `CAG`×19/`CAG`×41 disease-associated genotype. (The software used to generate graph (a) assigns the reference allele to non-calls.) `[HTT-400-both.png]`

Genotype graphs have a useful advantage over sequence graphs: they need not be constructed from contiguous sequence. We are free to construct a genotype graph from disjoint segments, such as exons in the case of whole exome sequencing (WES) data. We can exploit this advantage to study selected segments across the genome with limited computing power. We now turn to the definition of frequented regions and their associated quantities.

A path in a pangenome graph *supports* an FR if it satisfies two qualifying conditions based on two parameters, which deal with portions of the given path, termed *subpaths*:

> ***α*** sets the minimum fraction of nodes in the FR’s node cluster that a path’s *supporting subpath* must traverse; it ranges from *ϵ* to 1 (where *ϵ* is an arbitrarily small positive number to ensure that at least one node in the cluster is traversed by a supporting subpath). One can alternatively call 1 − *α* “the maximum fraction of *missing* FR nodes on a subpath.”
>
> ***κ*** sets the maximum number of consecutive nodes along a subpath that are *not* in the FR’s node cluster; it varies from 0 to ∞. When *κ* = 0, the subpath must traverse its FR nodes consecutively. One can call *κ* “the maximum number of consecutive *inserted* nodes on a subpath.”

In addition, qualifying subpaths must begin and end on FR nodes and be *maximal* : not part of a larger qualifying subpath. It is common for a path’s support to be greater than 1 because it contains distinct supporting subpaths. With *α* = 1, *κ* = 0, an FR is supported only by individuals that are genomically identical across the span of the FRs nodes (at the measured loci). For other values of *α* and *κ*, supporting paths may lack some FR nodes or contain nodes not belonging to the FR.

We note that FR analysis is sensitive to presence/absence, such as the case with indels. An insertion results in an extra node traversed by the individual’s path; a deletion results in lack of a node traversed by the individual’s path. These path variations lead to variations in FR support, depending on *α* and *κ*.

A few other terms: the *size* of an FR is the number of nodes in its cluster; the *total support S* of an FR is the total number of supporting subpaths. One can require that interesting FRs satisfy *minimum size* and/or *minimum support*. Figure 2 demonstrates a path and the variation of its support of an FR with different *α* and *κ* values.

**Figure 2:**
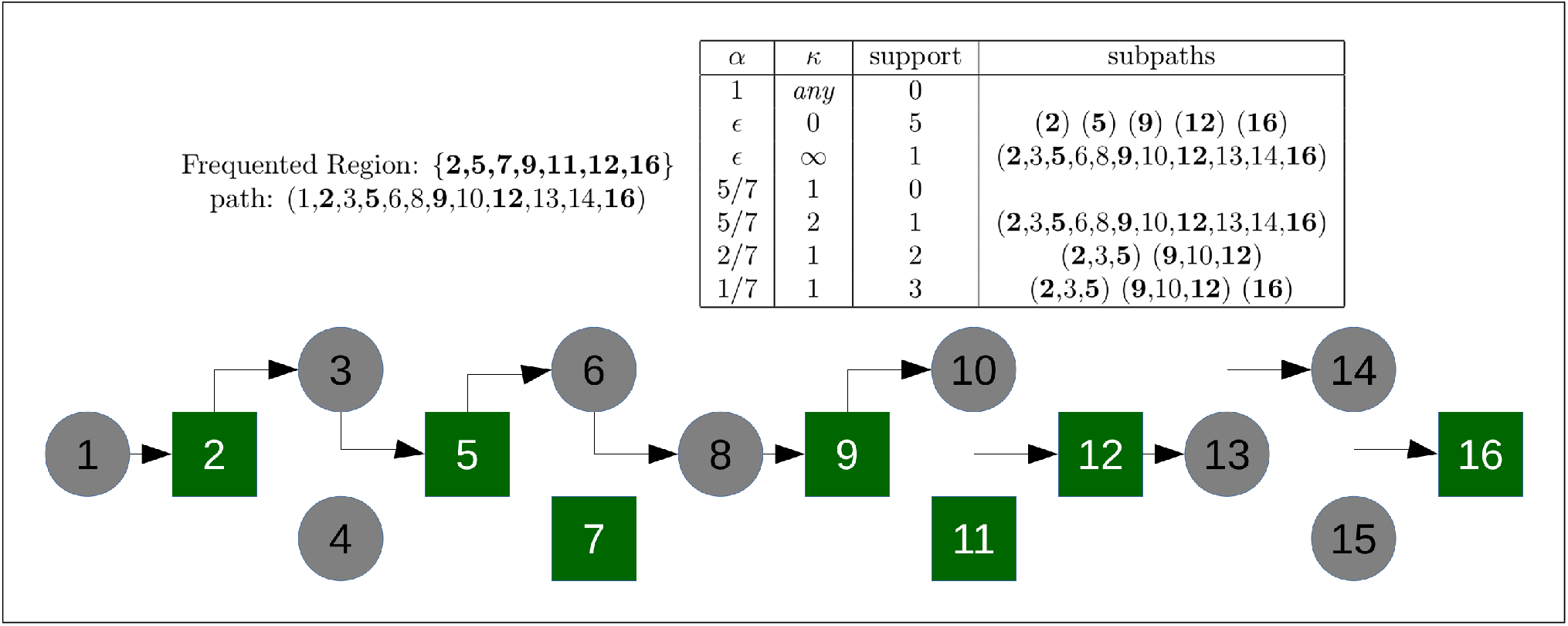
Dependence of one path’s support of a frequented region (FR) on parameters *α* and *κ*. Nodes aligned vertically represent variants at the same genomic location. This graph contains eight non-variant nodes (1, 2, 5, 8, 9, 12, 13, 16) and four bi-allelic variants (3/4, 6/7, 10/11, 14/15). The FR is designated by its node cluster {2,5,7,9,11,12,16}, shown with green rectangles; it has size 7. Supporting subpaths must begin and end on FR nodes, and must be *maximal* – not part of a larger supporting subpath. ***α* = 1**: all FR nodes must be traversed by a supporting subpath; the path contains only 5 of the 7 FR nodes, so support=0. ***α* = *ϵ, κ* = 0**: any single or consecutive FR nodes on the path support the FR; support=5. ***α* = *ϵ, κ* = ∞**: all FR nodes on the path span a single supporting subpath; support=1. ***α* = 5*/*7, *κ* = 1**: a supporting subpath must traverse 5 or more of the FR nodes; but *κ* = 1 requires that there be no more than one non-FR node *inserted* between traversed FR nodes; support=0. ***α* = 5*/*7, *κ* = 2**: a supporting subpath may have up to two nodes inserted between traversed FR nodes, so support=1. ***α* = 2*/*7, *κ* = 1**: a supporting subpath must traverse 2 or more FR nodes with up to one node inserted between FR nodes; support=2. ***α* = 1*/*7, *κ* = 1**: a supporting subpath must traverse 1 or more FR nodes with up to one node inserted between FR nodes, giving support=3. `[path-support.png]`

On the graphs studied here, each path is labeled “case” or “control”, corresponding to the individual’s status in a disease study. We separately tally an FR’s case and control subpath support, *S*_*case*_ and *S*_*ctrl*_, and prioritize FRs on case vs. control association using the support-based odds ratio

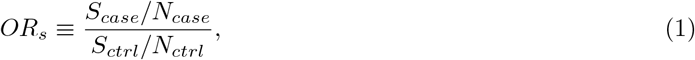

where *N*_*case*_ and *N*_*ctrl*_ are the number of case and control paths. The FR’s priority, an integer in our implementation, is defined by

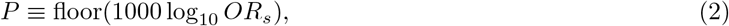

setting *P* = ± 2000 for *S*_*ctrl*_ = 0 or *S*_*case*_ = 0. (We use an integer for compatibility with integer-based priorities like total support.) To break ties, we favor larger total support, followed by smaller size. We can prioritize control-enhanced FRs by reversing the sign of *P* or, as we do here, prioritize case- and control-enhanced FRs equally by using |*P*|.

The problem of finding FRs is *computationally hard* : it is *#P-complete*[24], meaning it is not computationally feasible to find all FRs even for small data sets. However, we can identify FRs that are useful for our classification task using, for example, hierarchical clustering[25]. Identifying such clusters on clinical-scale data, however, is an *extreme clustering task* [26], requiring powerful computational resources. In order to analyze large regions one must implement an FR search algorithm that runs in polynomial rather than exponential time[27]. Cleary *et al*.[16] describe a heuristic algorithm using bottom-up, agglomerative clustering[25]. For the purpose of showing the efficacy of an FR-based approach for disease classification, and to gain insight into the algorithm specialization that will be necessary to perform this task at scale, we used a brute-force search for the results presented here, with some computation-reducing additions described in the examples. Our implementation^1^ has many optional parameters to control the search. The work was performed on a 128-CPU, 1-TB machine running Java under Clear Linux.

The graphs studied here were built from variant calls against the human reference genome provided in VCF files, rather than directly from sequencing reads. This does not affect the purpose of this report, which is to demonstrate the efficacy of the methods described herein. We set our FR search routine to leave no-call genotypes (denoted “./.” in the VCF) out of FRs; paths continue to traverse them and they are included in the support calculation. In the future, we plan to build graphs directly from whole-genome DNA sequencing reads, or reference-guided assemblies, which can reveal novel genomic content in the study population which impacts disease risk.

### Machine learning and supervised classification of disease status

Frequented regions, on their own, provide an informative way of viewing genomic variation that can be linked to disease. However, of more interest to the clinician is the ability to estimate genomic disease risk of a given individual. For this purpose, FRs provide a natural framework for machine learning, in particular supervised classification[16,21].

Supervised classification consists of *training* a classifier from a labeled training set, *validating* the classifier against a separate set of labeled data not used for training, and *testing* the classifier on unlabeled data. Classifiers operate on *feature vectors* which encode each sample’s quantitative association with each feature. In our work, those features are FRs and the associated quantity is path support: the feature vectors are *FR path support vectors*, one for each individual in a study.

Table 1 displays ten path support vectors for five FRs of a graph containing only four nodes. Classifying these data is an easy task, since case and control support are quite distinct. For complex diseases, association with disease status is much less clear – our thesis is that FRs provide useful classication results which are complementary to other methods.

**Table 1:** Ten support vectors from a four-node graph built from two neighboring variants, rs63750783 and rs334 on the *HBB* gene, taken from the 1000 Genomes Project Consortium (Nature 526, 68-74 (2015); doi:10.1038/nature15393), along with frequented region support quantities: case support *S*_*case*_, control support *S*_*control*_, support-based odds ratio *OR*_*s*_, and priority *P*. For demonstration purposes, we have assigned “case” status to the 137 individuals (5.5%) carrying the rs334 sickle cell disease allele. Nodes 1–4 represent, in order: rs63750783[C/C], rs63750783[C/T], rs334[T/T], and the disease-associated genotype rs334[T/A]. (Node 2 is a rare genotype carried by only two individuals.)

| sample | label | frequented region |  |  |  |  |
| --- | --- | --- | --- | --- | --- | --- |
|  |  | {1,4} | {4} | {1,3} | {3} | {1} |
| HG03559 | “case” | 1 | 1 | 0 | 0 | 1 |
| HG03558 | “case” | 1 | 1 | 0 | 0 | 1 |
| HG03571 | “case” | 1 | 1 | 0 | 0 | 1 |
| HG03578 | “case” | 1 | 1 | 0 | 0 | 1 |
| HG03577 | “case” | 1 | 1 | 0 | 0 | 1 |
| NA21128 | “control” | 0 | 0 | 0 | 1 | 0 |
| HG03965 | “control” | 0 | 0 | 0 | 1 | 0 |
| NA20845 | “control” | 0 | 0 | 1 | 1 | 1 |
| NA20846 | “control” | 0 | 0 | 1 | 1 | 1 |
| NA18523 | “control” | 0 | 0 | 1 | 1 | 1 |
| ... | ... | ... | ... | ... | ... | ... |
| | $S_{case}$ | 137 | 137 | 0 | 0 | 137 |
| | $S_{control}$ | 0 | 0 | 2365 | 2367 | 2365 |
| | $OR_s$ | $+\infty$ | $+\infty$ | $-\infty$ | $-\infty$ | 1.001 |
| | $P$ | +2000 | +2000 | -2000 | -2000 | 0 |

For comparison with our FR-based supervised classification, we perform a GWAS-type classification on *graph path traversal vectors* : vectors of 1’s and 0’s which denote presence/absence of each genotype, i.e. which nodes are/are not traversed by an individual’s path on the graph. This mode of classification is akin to standard GWAS, where each variant is treated independently. To assess the association of individual loci with the disease, we employ the standard measurement of *p*-value using the Cochran-Armitage test for trend, and use the term “GWAS-significant” for loci which have *p* < 5× 10^−8^ [28].

An important exercise in supervised learning is *k-fold cross-validation*.[29] One separates the individuals into *k* equally-sized groups and then uses each group to validate the classifier trained on the remaining individuals, until all individuals have been classified, and then summarizes the classification results. We ran 10-fold cross-validation with LIBSVM[30] as well as a number of binary classifiers from the Weka package[31]. Since LIBSVM outperformed the Weka classifiers in almost every case, we report only the LIBSVM results here. Cross-validations were run 10 times, using a different random number seed each time, to generate the reported mean values and variance of the classification results.

### Data sources

The data and analyses presented in this report are based on study data downloaded from dbGaP^2^, for General Research Use under dbGaP accessions phs001071.v1.p1 (HD) and phs000473.v2.p2 (SCZ). We used data downloaded from the 1000 Genomes Project Consortium (1kG)[32] for the SCD example.

## RESULTS

The research presented here on frequented regions in case-control disease studies demonstrates the efficacy of the method. Our brute-force algorithm allowed us to analyze graphs with up to around 1000 nodes on the available equipment. We now present results for three examples: sickle-cell disease (SCD), Huntington disease (HD), and schizophrenia (SCZ).

### Example 1: Sickle-cell disease (SCD)

Our first example is of a simple Mendelian disease: sickle-cell disease (SCD), an autosomal dominant disease associated with a SNP, rs334, on the *hemoglobin subunit beta* (*HBB*) gene on Chr 11.[33] Not having access to a case-control study of SCD with genomic data, we found that 137 of the 2,504 individuals in the 1kG database carry the deleterious allele. Since phenotype data is not provided, for the sake of demonstration we labeled those 137 individuals as “case” and labeled the other 2,367 individuals as “control”.

A pangenome graph provides information similar to, but more extensive than, a GWAS Manhattan plot. Figure 3 compares a GWAS Manhattan plot across the *HBB* gene with the genotype graph, for the full cohort of 2,504 subjects. One can see the associative genotypes in both cases, while the genotype graph provides a more extensive visualization of the variation amongst individuals. For explanatory purposes, we now discuss the meaning of certain choices of *α* and *κ* when searching for FRs on this graph.

**Figure 3:**
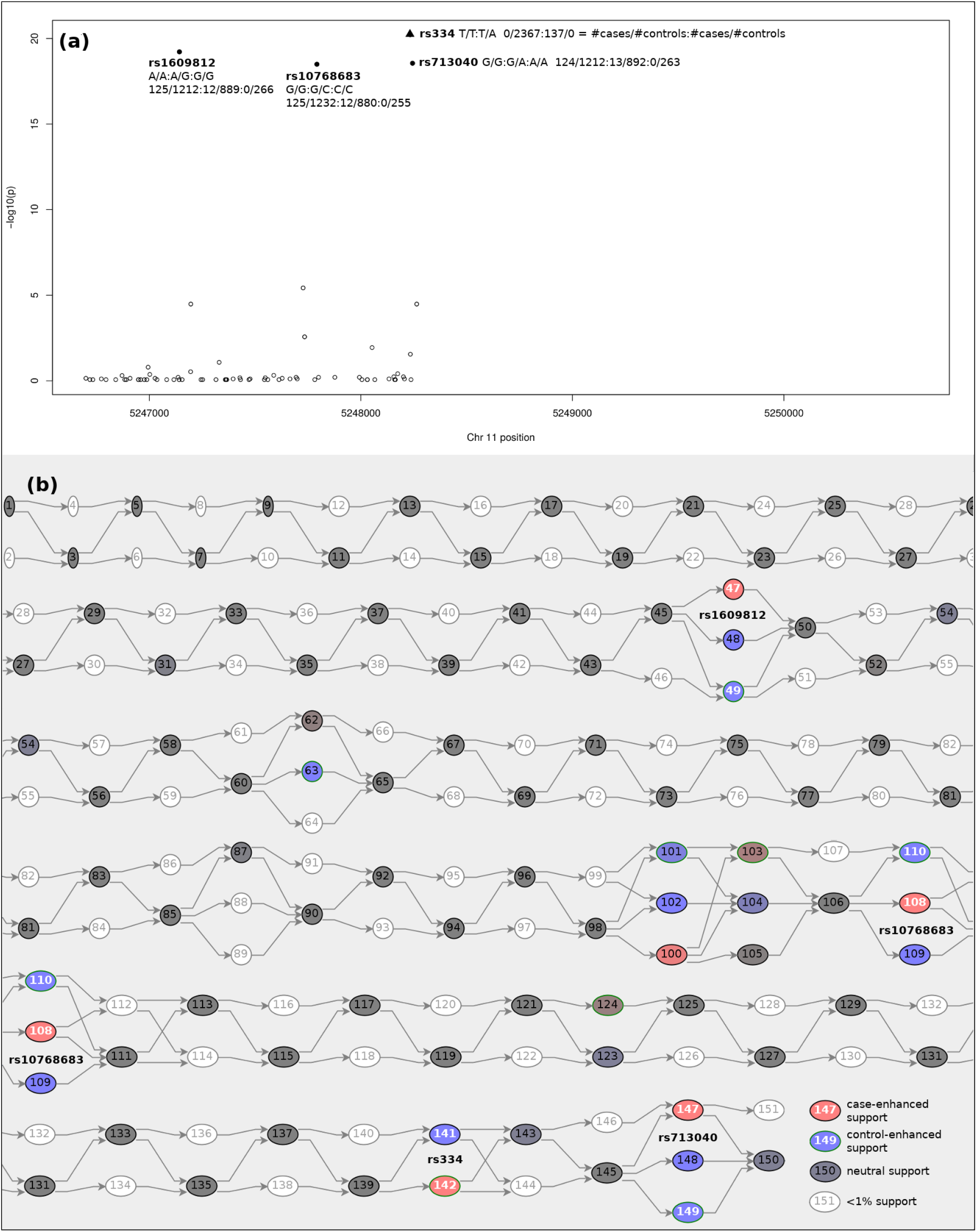
Comparison of GWAS and genotype graph analysis of the HBB gene for the 2504 individuals in the 1000 Genomes Consortium Project (Auton et al., *Nature* **526**, 68–74 (2015)). **(a)** GWAS Manhattan plot of the variants called against reference genome GRCh37, limited in the assay to the downstream half of this gene; **(b)** genotype graph constructed from those variants. *p*-values were calculated using the Cochran-Armitage test for trend. 137 individuals carry the A allele of the rs334 variant and have been labeled “case” for demonstration purposes. (The rs334 genotype, by design, has *p* = 0 in the GWAS analysis, and is placed for visibility at the top of the GWAS plot.) Both the GWAS Manhattan plot and pangenome graph display additional variants associated with these individuals: rs1609812, rs10768683, and rs713040. (Note that nodes on the right side of **(b)** are repeated below on the left side for clarity.) `[HBB-both.png]`

With *α* = 1 and *κ* = 0, supporting paths must traverse every FR node consecutively, without any inserted nodes; that is, with *α* = 1, *κ* = 0 an FR is supported only by individuals that are genomically identical across the span of the FR’s nodes (at the measured loci). As a result, path support for *α* = 1, *κ* = 0 tends to be low for FRs of large size.

The two nodes in Figure 3 corresponding to rs334 are 141 (`T/T`) and 142 (the deleterious `T/A`). The *single-node* FRs {141} and {142} therefore have complementary support and, by design, log_10_ *OR*_*s*_ = ±∞. Similarly, the single-node FRs {47}, {48}, and {49}, which correspond to the `A/A`, `G/G`, and `G/A` genotypes of GWAS-significant rs1609812, have case/control support of 125/1212, 0/266, and 12/889, or log_10_ *OR*_*s*_ = 0.25, −∞, and − 0.63. These five FRs would all be deemed interesting since they have *P* = ±2000, 250 and − 630. (It is worth noting that only 266 individuals, or 10.6%, all controls, carry the homozygous reference genotype, reminding us that the human reference genome often represents a minority genotype in a diverse population like 1kG[34].)

We note that with *α* = 1, *κ* = 0, *single-node* FR analysis is similar to GWAS, where each genotype is treated independently. An aspect of *α* = 1, *κ* = 0 is that path support decreases when one views larger FRs. For example, the case-associated FR {142,143,145,147} has support 121 rather than 137.

In order to find FRs which reveal multi-genotypic associations with non-shared intermediate nodes we set *κ* > 0. In fact, setting *κ* = ∞ is often desirable, since we are not concerned with the distance on the graph between nodes but, rather, are interested in discovering polygenic associations, regardless of where the variants are located on the genome. In practical terms, increasing kappa allows for the identification of shared genotypes among samples, even if those genotypes are separated by non-shared genotypes. Capturing these shared but distant genotypes in an FR allows us to test them for phenotype interactions as a group, thereby uncovering phenotypic effects due to genotype interactions even if the individual genotypes do not have a detectable effect on phenotype.

Figure 3 displays three strongly associated loci for our case/control assignment besides rs334 which was used for splitting cases and controls: rs1609812 (nodes 47–49), rs10768683 (nodes 108–110) and rs713040 (nodes 147–149). The case-enhanced genotypes form the FR {47,108,147} which, with *α* = 1, *κ* = ∞, has case/control support 124/1205 and log_10_ *OR*_*s*_ = 0.25 or *P* = 250. All but 13 of the case paths traverse these three nodes and therefore support this FR. 137 case-labeled subjects is too small a number from which to draw conclusions, but this particular FR might motivate study of the collective impact of these three loci on SCD, although it is also possible that their significance comes from their linkage to rs334 rather than from a biological effect of the variants. (In fact, dbSNP and ClinVar[35] report rs1609812:A, rs10768683:G, and rs713040:G as *benign* or *likely benign* alleles associated with SCD and/or *β* thalassemia.) With *α* = 1, *κ* =∞, FRs are supported by paths which traverse *all* of the FR nodes, revealing polygenic disease associations.

In conclusion, for a monogenic single-allele Mendelian disease like SCD, frequented regions of genotype graphs provide an alternative visualization of variation as well as an opportunity to discover multi-genotypic associations. They do not offer a large gain over GWAS in this scenario; their advantages become apparent in the analysis of multi-allelic Mendelian diseases with structural variants like Huntington disease (Example 2), and, more importantly, complex polygenic diseases like schizophrenia (Example 3).

### Example 2: Huntington disease (HD)

Huntington disease, or Huntington’s chorea, is a fully penetrant neurodegenerative monogenic disease caused by a dominantly inherited `CAG` trinucleotide repeat extension in the *huntingtin* gene (*HTT*) on Chr 4 (dbSNP rs71180116, ClinVar Variation ID 31916).[36] Unaffected individuals have fewer than 36 `CAG` repeats at this locus on both chromosomes; individuals carrying 36–39 repeats may or may not be affected, but pass a 50% disease risk to their offspring; individuals carrying 40 or more repeats suffer from the disease. Thus, there are a variety of rs71180116 genotypes which do and do not lead to disease status, as shown in Figure 1(b). We employed the *NINDS Family-Based Whole-Genome Sequencing to Find Modifiers of Age of Onset in Huntington’s Disease* study, dbGaP accession phs001071.v1.p1, and used the provided variant calls against human reference GRCh37 along with separately assayed `CAG` repeat lengths provided by the study authors. We labeled 27 case and 10 control subjects based on the reported phenotype and/or the length of the longer `CAG` repeat. All case individuals were heterozygous for the damaging allele. We limited the span of analysis to the first 400 bp of *HTT*, which includes rs71180116.

GWAS, often based on SNP array data or SNP calls using WGS or WES reads, are not typically designed to study structural variations such as this. Genotype graphs, conversely, make no distinction between simple variants like SNPs and larger variants: a node may contain any size genotype.

To enable analysis of a multi-allelic disease like HD, we set *α* < 1 to enable FR support by paths which traverse different nodes at the same locus. Table 2 displays the highest case- and control-supported FRs on the graph in Figure 1(b) as *α* is decreased. The graph has 6 control and 7 case nodes at the rs71180116 locus. When *α* ≤1*/*6, the FR {4,9,10,12,13,14}, containing all of the non-disease genotypes, is supported by all of the control paths; when *α* ≤ 1*/*7, the FR {3,5,6,7,8,11,15}, containing all of the disease-associated genotypes, is supported by all of the case paths. Decreasing *α* reveals multi-allelic FRs.

**Table 2:** Freqented Regions (FRs) with the largest case support *S*_*case*_ and control support *S*_*ctrl*_ as *α* is varied with *κ* = 0 throughout. The graph, shown in Figure 1(b), was built from the first 400 bases of the *HTT* gene, using variant calls against GRCh37 for 27 case and 10 control subjects in a Huntington disease study (dbGaP accession phs000473.v2.p2). Ties are broken by smaller size. Bold nodes represent genotypes of the disease-associated rs71180116 locus, and FRs composed of only those nodes appear when *α* ≤ 1*/*7 for cases and *α* ≤ 1*/*6 for controls.

| $\alpha$ | frequented region | size | $S_{case}$ | $S_{ctrl}$ |
| --- | --- | --- | --- | --- |
| 7/7 | {**3**} | 1 | 8 | 0 |
| 6/7 | {1,**3**,**8**,16,19,22,25} | 7 | 10 | 0 |
|  | {**3**,**6**,16,19,22,25,28} | 7 | 10 | 0 |
| 5/7 | {**3**,**6**,**7**,16,19,22,25} | 7 | 15 | 0 |
|  | {**3**,**6**,**8**,16,19,22,25} | 7 | 15 | 0 |
|  | {**3**,**7**,**8**,16,19,22,25} | 7 | 15 | 0 |
| 4/7 | {**3**,**6**,**7**,**8**,16,19,22} | 7 | 19 | 0 |
| 3/7 | {**3**,**5**,**6**,**7**,**8**,**15**,16,19,22} | 9 | 22 | 0 |
| 2/7 | {**3**,**5**,**6**,**7**,**8**,**11**,**15**,16,19,22} | 10 | 23 | 0 |
| 1/7 | {1,**3**,**5**,**6**,**7**,**8**,**11**,**15**,16,26,29} | 11 | 29 | 0 |
|  | {1,2,**3**,**5**,**6**,**7**,**8**,**11**,**15**,26,29} | 11 | 29 | 0 |
|  | {**3**,**5**,**6**,**7**,**8**,**11**,**15**} | 7 | 27 | 0 |
| 6/6 | {**4**} | 1 | 0 | 3 |
| 5/6 | {**4**,**10**,16,19,22,25} | 6 | 0 | 5 |
|  | {1,**4**,**10**,16,19,22} | 6 | 0 | 5 |
| 4/6 | {1,**4**,**10**,**12**,16,19} | 6 | 0 | 6 |
|  | {1,**4**,**10**,**13**,16,19} | 6 | 0 | 6 |
|  | {1,**4**,**10**,**14**,16,19} | 6 | 0 | 6 |
|  | {**4**,**10**,**13**,16,19,22} | 6 | 0 | 6 |
|  | {**4**,**10**,**14**,16,19,22} | 6 | 0 | 6 |
| 3/6 | {1,**4**,**10**,**12**,**13**,**14**,16,19} | 8 | 0 | 8 |
| 2/6 | {1,**4**,**9**,**10**,**12**} | 5 | 0 | 8 |
| 1/6 | {1,**4**,**9**,**10**,**12**,**13**,**14**,25,26,31,32} | 11 | 0 | 12 |
|  | {**4**,**9**,**10**,**12**,**13**,**14**} | 6 | 0 | 10 |

We applied 10-fold cross-validation to the path support vectors for 692 FRs generated with *α* = *E, κ* = 0. LIBSVM consistently delivered perfect cross-validation. This is unsurprising, given the association of the HD genotypes with disease status. The final example, exploring a case-control schizophrenia study, exhibits imperfect classification which improves when the classifier is trained on FR path support rather than graph path traversal.

### Example 3: Schizophrenia (SCZ)

Schizophrenia is a heritable psychiatric disorder that affects up to 1% of the general population.[37,38] Large-scale GWAS point to a large number of genes contributing to its pathophysiology, due to both rare and common variants.[39–42] A composite picture of heritability remains elusive.

In this example, we explore the application of frequented regions to a case-control study of 12,380 Swedish individuals, *Sweden-Schizophrenia Population-Based Case-Control Exome Sequencing*, dbGaP study accession phs000473.v2.p2.[41] We used the variant calls against GRCh37 provided by the study.

First, we removed subjects diagnosed with bipolar disorder rather than SCZ, leaving 11,209 individuals. Then, we removed randomly selected control-labeled subjects to stratify the cohort at 4,966 cases and 4,966 controls, since balanced designs are known to provide better supervised classification[43].

With our algorithm and computational power, we must explore limited genomic segments. The most strongly disease-associated segment is the HLA region of Chr 6, shown in Figure 4, from which we selected the *HLA-A, HLA-B*, and *HLA-C* genes to build graphs labeled HLAA, HLAB and HLAC, with 1,166, 1,291, and 1,091 nodes, respectively. For comparison, we selected two low-association segments: one on Chr 6:151627034–151939181 for a graph labeled SCZ6A with 1,093 nodes, and one on Chr 14:31349968–31647448 for a graph labeled SCZ14C with 1,084 nodes. GWAS Manhattan plots of these five segements are shown in Figure 5.

**Figure 4:**
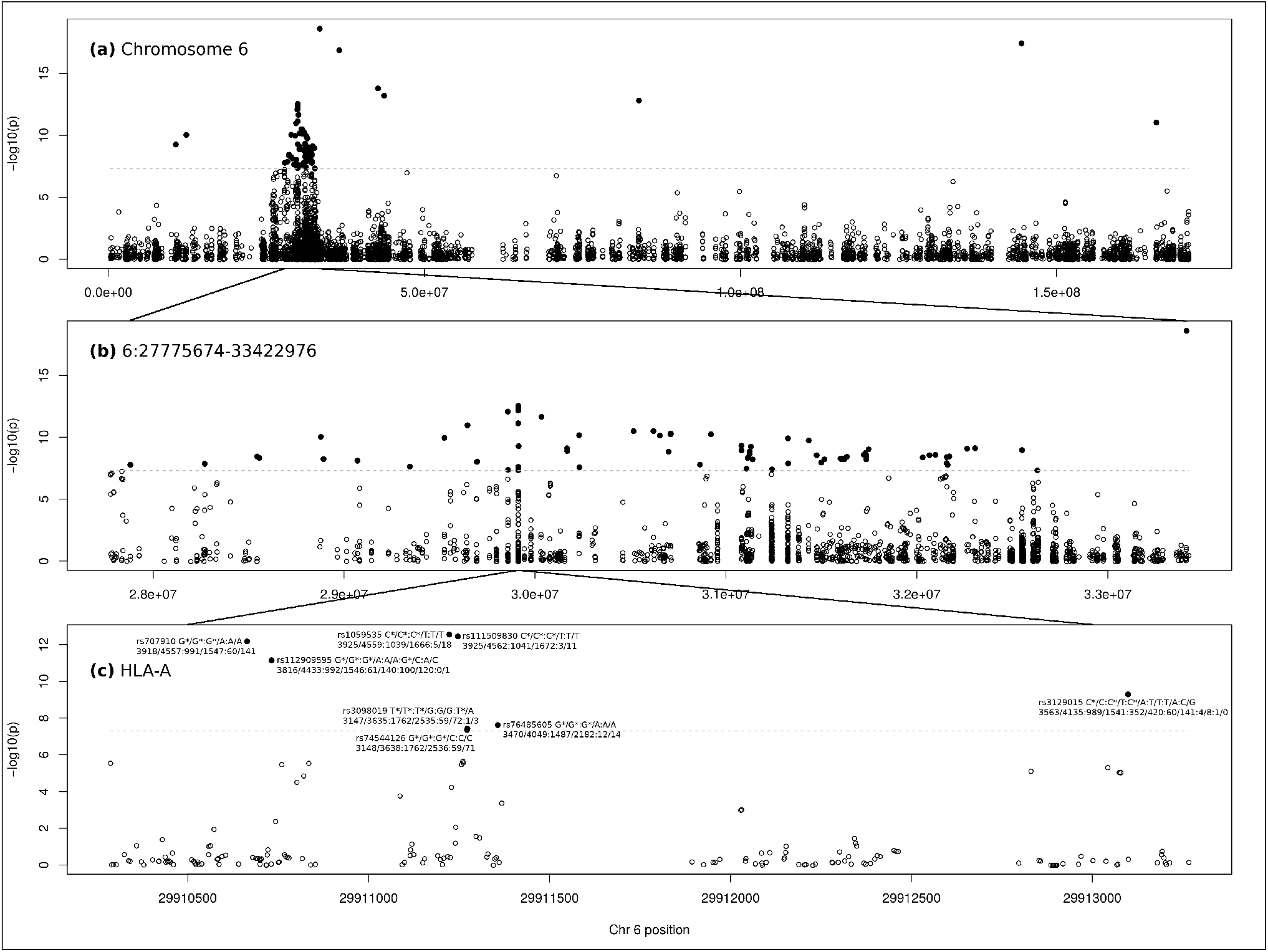
Manhattan plots of the variants called against reference genome GRCh37 Chr 6 in an exome sequencing study of schizophrenia amongst Swedes (dbGaP accession phs000473.v2.p2). **(a)** Chromosome 6; **(b)** a span of high case vs. control association on Chr 6, slighly larger than the HLA region; **(c)** the *HLA-A* gene, which carries eight significant (*p* < 5 × 10^−8^) variants (two of which form a two-base haplotype). *p*-values were calculated using the Cochran-Armitage test for trend. `[6-GWAS.png]`

**Figure 5:**
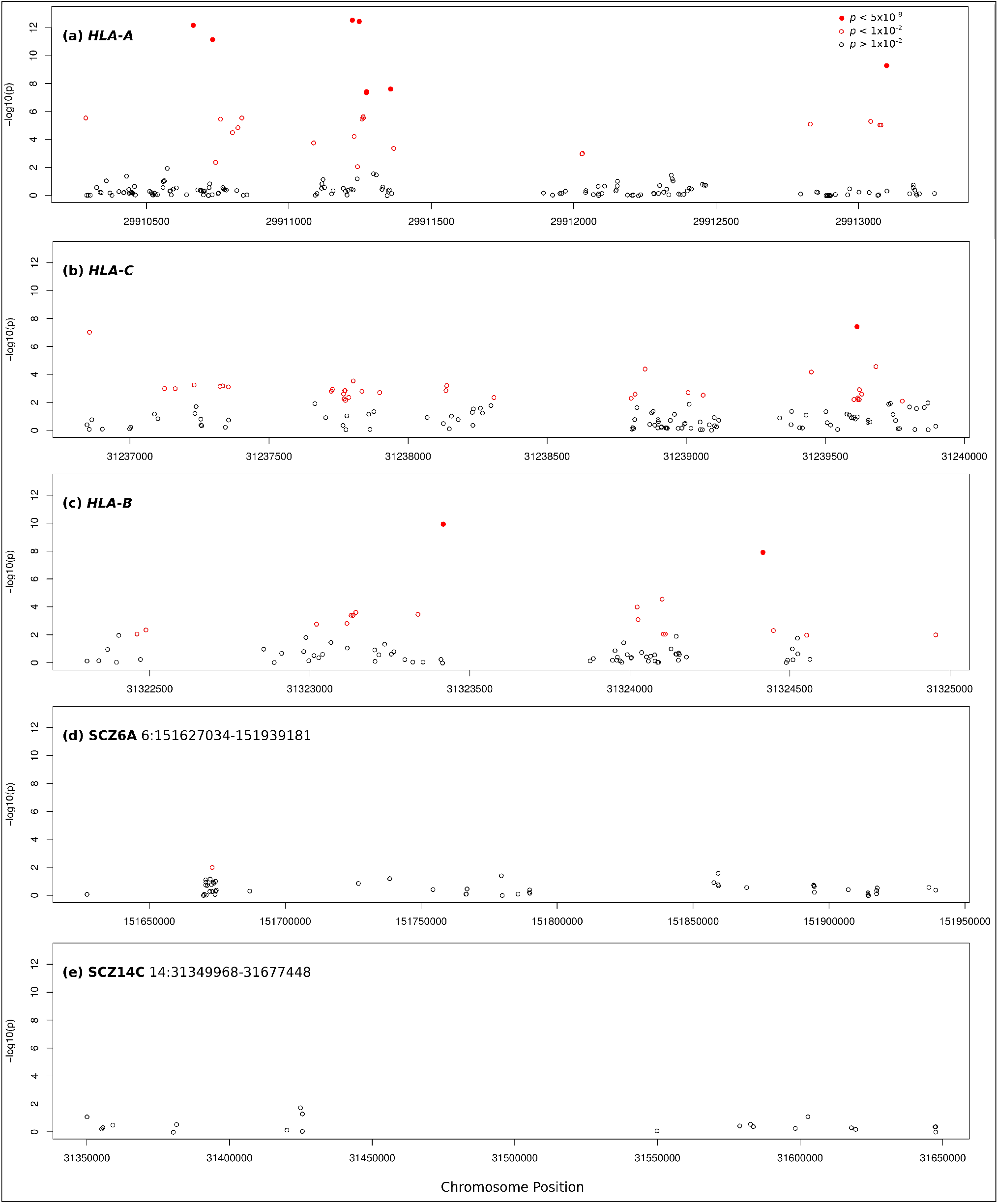
Manhattan plots of the five segments used for frequented region analysis, from variants called against reference genome GRCh37 in an exome sequencing study of schizophrenia amongst Swedes (dbGaP accession phs000473.v2.p2). **(a)** the *HLA-A* gene; **(b)** the *HLA-C* gene; **(c)** the *HLA-B* gene; **(d)** a segment on Chr 6 labeled SCZ6A; **(e)** a segment on Chr 14 labeled SCZ14C. *p*-values were calculated using the Cochran-Armitage test for trend. `[ALL-GWAS.png]`

The goal of this example is to determine whether supervised classification is improved when we train a classifier on FR path support rather than graph path traversal (GWAS). FRs bring out disease association with clusters of loci. For example, no locus in the SCZ14C range has a *p*-value less than 1× 10^−2^, yet many FRs, supported by a subset of paths, can be found that exhibit strong disease association. The combination of such FRs may provide improved classification. We now make a few points about frequented regions using the *HLA-A* gene, which contains eight GWAS-significant loci (Figure 4).

First, it is worth noting the effect of haplotypes on FR support. Figure 6(a) shows that two of the eight associating loci are strongly linked based on minimal changes to their support: rs74544126 and rs3098019 are on consecutive bases and form haplotype pairs: `GT/GT` and `GT/CG`. (With a graph built from assemblies or reads rather than SNP calls, these haplotypes and much larger genotypes will naturally appear.) The FR {784,787}, corresponding to `GT/CG`, has total support equal to one less than {784} alone. The presence of haplotypes across separate nodes increases an FR’s size with little effect on its support.

**Figure 6:**
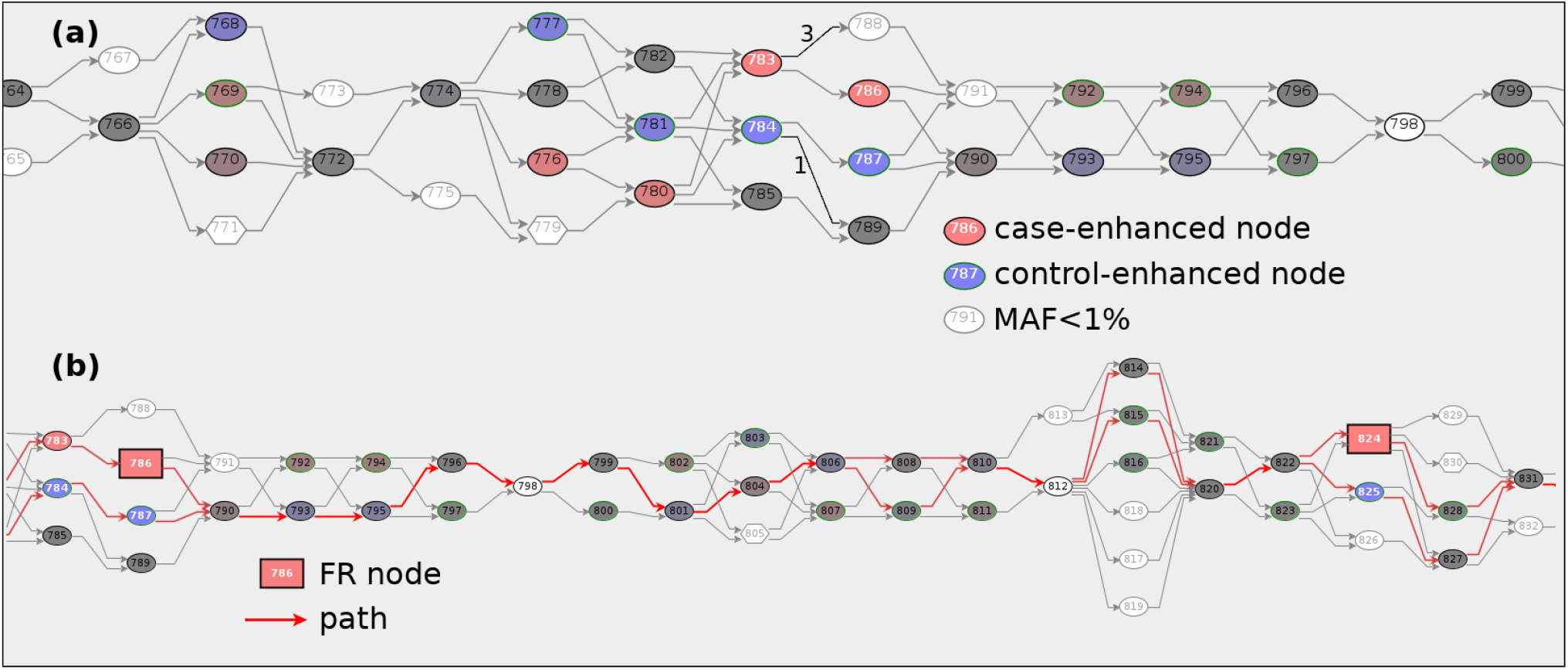
Portions of a graph of the *HLA-A* gene on Chr 6 built from variant calls for 9,932 individuals in an exome sequencing study of schizophrenia amongst Swedes (dbGaP accession phs000473.v2.p2). **(a)** Two pairs of tightly-linked neighboring nodes: 783,786 and 784,787. All but four paths that traverse 783 or 784 also traverse 786 or 787, respectively. The pairs represent two-base haplotypes. **(b)** The frequented region {786,824}. Two case-labeled paths demonstrate how support diminishes when nodes are added to an FR with *α* = 1. The single-node FR {786} has case+control support of 3,144+2,872=6,016; {824} has 3,468+3,147=6,615; while {786,824}, supported by paths which traverse nodes 786 *and* 824, has 2,737+2,502=5,239. If *α* is small, paths traversing nodes 786 *or* 824 support {786,824}, giving 3,875+3,557=7,432. `[HLAA-783-786-786-824.png]`

However, when *α* = 1, combination of less strongly-linked nodes reduces support. This is the case with the FR shown in Figure 6(b). While {786} has *S* = 6016 and {824} has *S* = 6655, the combined FR {786,824 has *S* = 5239. When *α* = 1, combining nodes generally *decreases* support. When *α* is small, however, paths traversing {786} *or* {824} will support {786,824}, resulting in *S* = 7482. With small *α*, combining nodes *increases* support.

The eight GWAS-significant variants on *HLA-A* shown in Figure 4 provide eight case-enhanced nodes on the HLAA graph. All are shared by many affected and unaffected individuals: with *α* = 1, 49.3% of the case paths and 44.5% of the control paths support the full eight-node FR, giving *OR*_*s*_ = 1.106. With *α* = 1 we emphasize fully shared genotypes, i.e. the *intersection* of individuals’ genotypes. But complex diseases are associated with a wide array of genotypes, of which a subset are carried by any particular individual. Therefore, *small α* is valuable in revealing FRs that represent the *union* of individuals’ genotypes.

Repeating the above with *α* = *ϵ* to determine support by paths that contain at least *one* of the eight genotypes results in support by 79.1% of the case paths and 72.9% of the control paths. Since FRs with small *α* are supported by a larger number of paths, they provide more extensive input to a supervised learning classifier.

For small *α* and *κ* =∞, a path will support an FR by either 0 or 1, since the maximal subpath that contains FR nodes is unique when any number of inserted nodes is allowed. One can think of *α* = *E, κ* = as a *binary test* of whether a path traverses any of the FR’s nodes. For small *α* and *κ* = 0, however, a path’s support will be nearly equal to the number of FR nodes that it traverses, reduced slightly when the path traverses two or more FR nodes consecutively. One can think of *α* = *E, κ* = 0 as providing an approximate *count of the FR nodes* that a path traverses. The same paths will provide support in both cases, but with *κ* = 0 the path support vectors will have a variety of support values, rather than 0 or 1 with *κ* = ∞, and this may impact supervised learning and classification. (A range of *α* and *κ* values may be scanned to find an optimal combination for classification; we have done that, and found that extreme values of *α* and *κ* are sufficient for our purposes here.)

To establish a baseline of supervised classification, we ran 10-fold cross-validation on the path traversal of each graph (GWAS), disregarding no-call nodes. Results are presented in Table 3. The best path traversal classification is found on HLAC with 54.5% of individuals called correctly, specificity=0.502, sensitivity=0.588, and Matthew’s Correlation Coefficient MCC=0.091. (A random classifier would call 50% correctly, with specificity=sensitivity=0.5 and MCC=0.) Our low-association segments, SCZ6A and SCZ14C, provide poorer path traversal classification, as one would expect. If frequented regions are to be a useful alternative to standard GWAS, they must perform better than this baseline.

**Table 3:** Results from 10-fold LIBSVM cross-validation of path traversal vectors (GWAS) and FR path support vectors from six graphs. Each cross-validation was run 10 times, varying random number seeds, to generate the shown mean and standard deviation. Graphs were built using variant calls from an exome sequencing study of schizophrenia amongst Swedes (dbGaP accession phs000473.v2.p2). The graphs labeled HLAA, HLAB, and HLAC were built from the genes *HLA-A, HLA-B*, and *HLA-C*; SCZ6A and SCZ14C are from weakly disease-associated segments on Chr 6 and 14; SCZ6A+SCZ14C is their combined graph. MCC is Matthew’s Correlation Coefficient. # indicates the number of analyzed graph nodes for path traversal classification and the number of analyzed frequented regions for FR path support classification.

| graph | feature vectors | # | correct | sensitivity | specificity | MCC |
| --- | --- | --- | --- | --- | --- | --- |
| HLAA | path traversal (GWAS) | 978 | 53.0±0.3% | 0.653±0.005 | 0.408±0.003 | 0.063±0.006 |
| | FR path support | 820 | 55.5±0.2% | 0.649±0.005 | 0.461±0.003 | 0.112±0.003 |
| HLAB | path traversal (GWAS) | 1038 | 53.9±0.3% | 0.544±0.004 | 0.534±0.003 | 0.079±0.005 |
| | FR path support | 975 | 57.2±0.2% | 0.559±0.003 | 0.586±0.003 | 0.145±0.005 |
| HLAC | path traversal (GWAS) | 896 | 54.5±0.4% | 0.502±0.004 | 0.588±0.005 | 0.091±0.007 |
| | FR path support | 810 | 56.1±0.1% | 0.550±0.002 | 0.572±0.002 | 0.122±0.003 |
| SCZ6A | path traversal (GWAS) | 1031 | 51.7±0.3% | 0.468±0.003 | 0.567±0.002 | 0.035±0.005 |
| | FR path support | 629 | 55.2±0.2% | 0.541±0.003 | 0.564±0.005 | 0.105±0.003 |
| SCZ14C | path traversal (GWAS) | 986 | 51.9±0.2% | 0.418±0.004 | 0.619±0.004 | 0.038±0.005 |
| | FR path support | 551 | 54.3±0.3% | 0.464±0.004 | 0.622±0.003 | 0.087±0.006 |
| SCZ6A+SCZ14C | path traversal (GWAS) | 2168 | 52.4±0.1% | 0.485±0.002 | 0.564±0.003 | 0.049±0.003 |
| | FR path support | 1173 | 59.5±0.2% | 0.570±0.002 | 0.620±0.003 | 0.190±0.003 |

We then searched for FRs on each graph, requiring *S* ≥ 100 and|*P*| ≥ 1, which is comparable to GWAS requiring MAF>1% and log10 *OR* > 0.001. This is too large a problem for our algorithm without further reduction. To make the problem feasible, we added three constraints to the FR search routine:

(1) Interesting FRs must contain a specified *starter node* (`FRFinder` parameter `--requirednodes`).

(2) Interesting FRs must have higher priority than those previously found or be a subset of a previously found FR with the same priority (`--keep=subset`).

(3) Interesting FRs in step *n* must contain the nodes in the FR found in step *n*−1 (`--requirebestnodeset`).

(1) and (2) do not incur a loss of generality since we run every node as a starter node, and supersets of an FR with the same priority are generally uninteresting. Constraint (3), however, which extends the most interesting FR one node at a time, greatly shrinks the available space of FRs. We found that (3) still provides a useful set for classification. (Note that a node will generally appear in many more FR runs than the one in which it is the starter.) For each starter node, we keep the best (last and largest) FR found for training of a supervised classifier.

With those constraints in place, we set *α* = *E, κ* =∞ and searched for FRs of each graph. The resulting 10-fold cross-validations are shown in Table 3 and Figure 7. Supervised learning of FR path support consistently provided better classification than that of path traversal. Of particular note is the improvement on SCZ6A, from 51.7% to 55.2% correct calls. *A segment with no significantly associated loci provided better classification using frequented regions than even highly-associated segments did using path traversal*.

**Figure 7:**
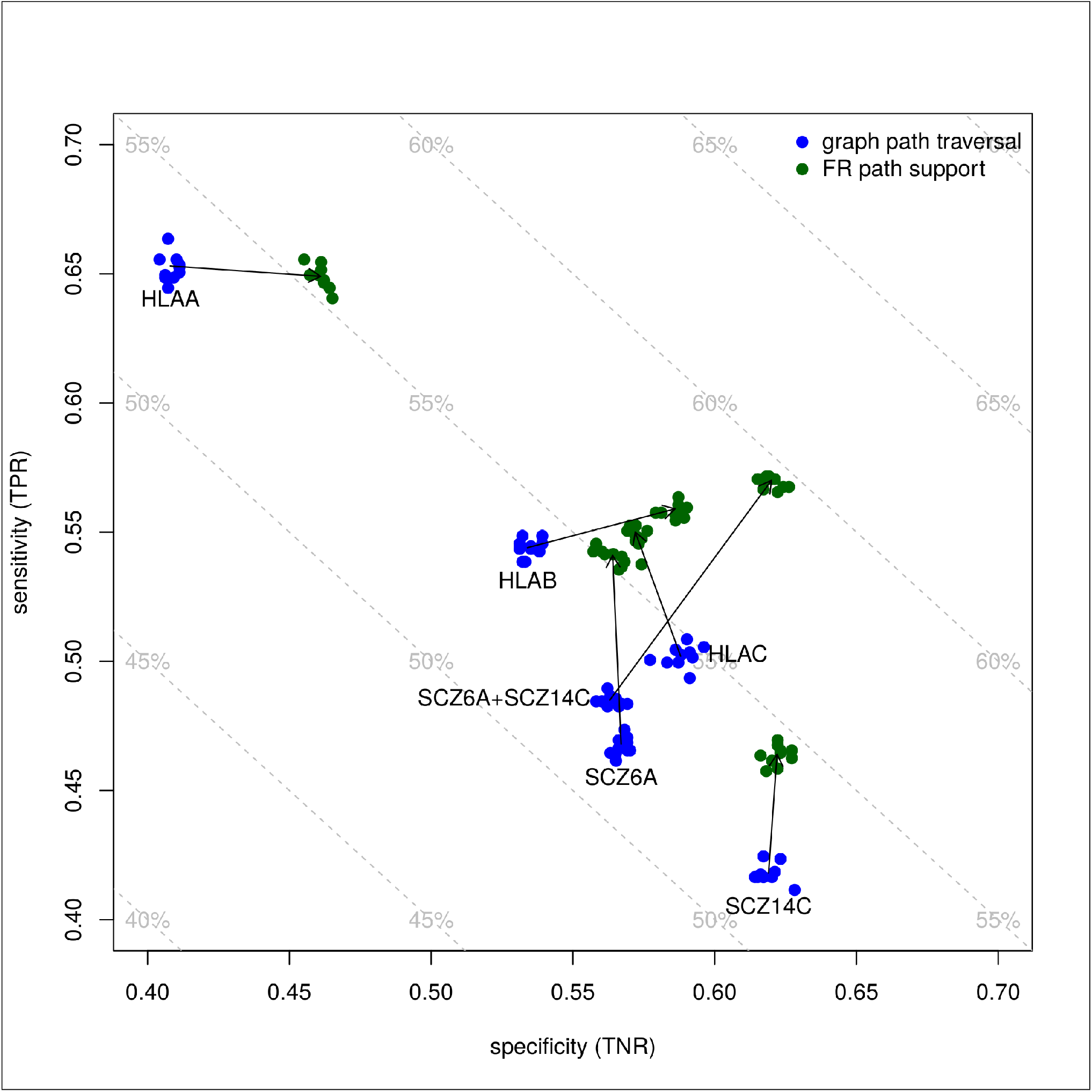
Supervised classification results for feature vectors built from graph path traversal (GWAS) (blue) and frequented region path support (green). Six graphs were studied: three from the highly disease-associated genes *HLA-A, HLA-B*, and *HLA-C*, two from low-association segments 6:151627034–151939181 (SCZ6A) and 14:31349968–31647448 (SCZ14C), and the combination of SCZ6A and SCZ14C. LIBSVM 10-fold cross-validation was run 10 times for each dataset, varying the random number seed for each run. Arrows start and end on mean values; dashed gray lines indicate total correct classification. `[sensitivity-specificity.png]`

Finally, in order to explore the improvement that occurs when combining low-association segments, we built a graph from SCZ6A and SCZ14C. This graph has 2,168 nodes and led to 1,173 interesting FRs. The classification results were surprising: while path traversal classification resulted in 52.4% correct calls (only slightly better than SCZ6A and SCZ14C individually), classification with frequented regions resulted in 59.5% correct calls and MCC=0.190, both higher than any of the other runs. Figure 7 shows how sensitivity and specificity improved equally, leading to a large increase in total correct calls. This particular result encourages the pursuit of frequented regions composed of intervals across the entire genome.

Although, overall, the classification gains are not huge, it is clear that, on this SCZ dataset, frequented regions provide a better basis for supervised learning than do individual genotypes.

Twin studies provide us with an estimate of the maximum achievable classification of SCZ. Since there are many identical twins for which one sibling is affected and the other is not, we know that perfect classification is impossible even if we were to use the entire genome as input, since environmental and other non-genomic factors affect disease status. The probandwise concordance rates for several studies of identical twins are reported in [44] and cluster around 45%. If one had a cohort of 100 twins, for which one or both siblings is affected by SCZ, along with a cohort of 200 control individuals and a perfect classifier that classifies all SCZ genomes correctly, one would incorrectly classify 55 individuals as SCZ-positive, for a FPR=55/255=0.22, with 86% total correct calls and specificity=0.78.

We conducted a graph path traversal (GWAS) analysis of the 307 GWAS-significant loci on autosomal chromosomes in the Swedish SCZ study (891 total nodes). LIBSVM classified 60.1 ± 0.2% of a 4969/4969 cohort correctly, with sensitivity=0.493± 0.002, specificity=0.708 ± 0.002, and MCC=0.206 ± 0.004. Given the results presented here, we expect that a genome-wide analysis of frequented regions would add at least 2–5% to this 60% correct classification, and perhaps much more.

## DISCUSSION

This report presents a proof of principle for using frequented regions of genotype graphs in the study of complex diseases. The sickle-cell disease and Huntington disease examples help to explain the methodology in a simplified setting, while the Swedish schizophrenia study demonstrates improvement in genomic classification of a complex disease. The SCZ example employed small graphs built from exonic variants, mostly SNPs, which are thought to capture only 23% of SCZ susceptibility[40]. We believe that larger graphs, built directly from whole genome sequencing reads, without reliance on the human reference genome, will provide much better classification. In any case, our preliminary results suggest that frequented regions are worthy of further study.

We are not the first to apply machine learning to SCZ: a recent study[45] applied ML to feature vectors composed of SNP calls from the same Swedish study as used here, on 50 genes chosen *a priori* based on bioinformatic criteria. In particular, the authors focused on rare (predicted functional) variants, with MAF≤ 1%. They obtained remarkably good classification results from a selected 2545/2545 cohort. Our approach differs greatly from theirs: our method emphasizes the *discovery* of associated *collections* of genotypes, without regard to rate of their occurrence in the general population, including both common and rare alleles. In fact, ours is a standalone analysis which uses as input solely the pangenomic graph of the study individuals’ genotypes.

It is important to emphasize that our classification and cross-validation operated on individuals that were members of a relatively non-diverse population: ethnic Swedes. It is unlikely to perform well, as trained, on genomes from other populations. In fact, we believe that this is a fundamental aspect of genomic diagnosis of complex diseases: in order to find signatures that associate with disease status, one must have a sufficiently uniform genomic background. If FR analysis is to result in a clinical diagnostic, that diagnostic will have to be tuned for the population of the individuals being tested, based on ethnicity or perhaps a distinguishing set of markers. This is not a statistical problem for schizophrenia, which is common in all human populations. It becomes a problem, however, for rare diseases, when statistical power may be insufficient within a particular population.

Another challenge with classification based on FRs is the task of applying the classifier to an individual that is not in the cohort that was used to build the graph. In principle, if the graph is comprehensive enough, such an individual will have a fully-defined path for which the FR support vectors may be computed. However, there is no guarantee that an individual being tested does not have a relevant genotype that is missing from the training cohort.

We hope to find FRs on much larger graphs built from WGS reads, without involvement of a reference genome, and to measure their effectiveness in disease classification and other tasks. We have the long-term goal of building an FR-based disease classification *appliance*, which would be trained for a particular disease and population, and then used to test unlabeled genomes from that population. This sort of device could be useful in applications such as treatment, where, say, the successful medications for affected individuals would be used to label training data in order to suggest a medication to try with new patients. Case-control studies are only one of many types of study that can be analyzed with frequented regions.

That pursuit will likely find new and important disease-associated genotypes that are not present in the human reference genome. We believe that new pangenomic analyses like frequented regions of genotype graphs are an extremely important path for genomic disease research.

## Data Availability

The case-control data used are available from dbGaP and the 1000 Genomes Consortium, the latter publicly available, the former requiring dbGaP approval. The dbGaP accessions used are phs001071.v1.p1 and phs000473.v2.p2. The code written and used is publicly available on GitHub under the repositories sammyjava/GWAS and sammyjava/pangenomics.

https://github.com/sammyjava/GWAS

https://github.com/sammyjava/pangenomics

https://www.ncbi.nlm.nih.gov/projects/gap/cgi-bin/study.cgi?study_id=phs001071.v1.p1

https://www.ncbi.nlm.nih.gov/projects/gap/cgi-bin/study.cgi?study_id=phs000473.v2.p2

https://www.internationalgenome.org/

## Acknowledgements

We thank Callum Bell and Andrew Farmer for insightful comments on drafts of the manuscript.

The *NINDS Family-Based Whole-Genome Sequencing to Find HD Modifiers* datasets used for the analysis described in this manuscript were obtained from dbGaP at http://www.ncbi.nlm.nih.gov/gap through dbGaP accession number phs001071.v1.p1. We thank the submitting investigator Steven Finkbeiner.

The *Sweden-Schizophrenia Population-Based Case-Control Exome Sequencing* datasets used for the analysis described in this manuscript were obtained from dbGaP at http://www.ncbi.nlm.nih.gov/gap through dbGaP accession number phs000473.v2.p2. Samples used for data analysis were provided by the Swedish Cohort Collection supported by the NIMH grant R01MH077139, the Sylvan C. Herman Foundation, the Stanley Medical Research Institute and The Swedish Research Council (grants 2009-4959 and 2011-4659). Support for the exome sequencing was provided by the NIMH Grand Opportunity grant RCMH089905, the Sylvan C. Herman Foundation, a grant from the Stanley Medical Research Institute and multiple gifts to the Stanley Center for Psychiatric Research at the Broad Institute of MIT and Harvard.

We also thank the International Genome Sample Resource for keeping the 1000 Genomes data freely available.

The work presented here was partly funded and computational resources provided by the National Center for Genome Resources, Santa Fe, New Mexico, USA. Partial funding also came from NSF award #1759522.

## Footnotes

1 `FRFinder` available at https://github.com/sammyjava/pangenomics

2 https://www.ncbi.nlm.nih.gov/gap/

## References

[1] E. A. Boyle, Y. I. Li, and J. K. Pritchard. An Expanded View of Complex Traits: From Polygenic to Omnigenic. Cell, 169(7):1177–1186, Jun 2017.

[2] E. Gnin. Missing heritability of complex diseases: case solved? Hum. Genet., 139(1):103–113, Jan 2020.

[3] A. R. Martin, M. J. Daly, E. B. Robinson, S. E. Hyman, and B. M. Neale. Predicting Polygenic Risk of Psychiatric Disorders. Biol. Psychiatry, 86(2):97–109, 07 2019.

[4] P. M. Visscher, M. A. Brown, M. I. McCarthy, and J. Yang. Five years of GWAS discovery. Am. J. Hum. Genet., 90(1):7–24, Jan 2012.

[5] Aude Saint Pierre and Emmanuelle Génin. How important are rare variants in common disease? Brief. Funct. Genomics, 13(5):353–361, September 2014.

[6] A. Torkamani, N. E. Wineinger, and E. J. Topol. The personal and clinical utility of polygenic risk scores. Nat. Rev. Genet., 19(9):581–590, 09 2018.

[7] Jason H Moore. The ubiquitous nature of epistasis in determining susceptibility to common human diseases. Hum. Hered., 56(1-3):73–82, 2003.

[8] M D Ritchie, L W Hahn, N Roodi, L R Bailey, W D Dupont, F F Parl, and J H Moore. Multifactor-dimensionality reduction reveals high-order interactions among estrogen-metabolism genes in sporadic breast cancer. Am. J. Hum. Genet., 69(1):138–147, July 2001.

[9] M. D. Ritchie and K. Van Steen. The search for gene-gene interactions in genome-wide association studies: challenges in abundance of methods, practical considerations, and biological interpretation. Ann Transl Med, 6(8):157, Apr 2018.

[10] Elena S Gusareva and Kristel Van Steen. Practical aspects of genome-wide association interaction analysis. Hum. Genet., 133(11):1343–1358, November 2014.

[11] Lorenzo Bomba, Klaudia Walter, and Nicole Soranzo. The impact of rare and low-frequency genetic variants in common disease. Genome Biol., 18(1):77, April 2017.

[12] Futao Zhang, Eric Boerwinkle, and Momiao Xiong. Epistasis analysis for quantitative traits by functional regression model. Genome Res., 24(6):989–998, June 2014.

[13] Rosanna Upstill-Goddard, Diana Eccles, Joerg Fliege, and Andrew Collins. Machine learning approaches for the discovery of gene-gene interactions in disease data. Brief. Bioinform., 14(2):251–260, March 2013.

[14] Sebastian Okser, Tapio Pahikkala, Antti Airola, Tapio Salakoski, Samuli Ripatti, and Tero Aittokallio. Regularized machine learning in the genetic prediction of complex traits. PLoS Genet., 10(11):e1004754, November 2014.

[15] Duc-Hau Le. Machine learning-based approaches for disease gene prediction. Brief. Funct. Genomics, June 2020.

[16] A. Cleary, T. Ramaraj, I. Kahanda, J. Mudge, and B. Mumey. Exploring Frequented Regions in Pan-Genomic Graphs. IEEE/ACM Trans Comput Biol Bioinform, 16(5):1424–1435, 2019.

[17] T. Marschall, M. Marz, et al. Computational pan-genomics: status, promises and challenges. Brief. Bioinformatics, 19(1):118–135, 01 2018.

[18] J. M. Eizenga, A. M. Novak, J. A. Sibbesen, S. Heumos, A. Ghaffaari, G. Hickey, X. Chang, J. D. Seaman, R. Rounthwaite, J. Ebler, M. Rautiainen, S. Garg, B. Paten, T. Marschall, J. Sirén, and E. Garrison. Pangenome Graphs. Annu Rev Genomics Hum Genet, 21:139–162, Aug 2020.

[19] Heng Li, Xiaowen Feng, and Chong Chu. The design and construction of reference pangenome graphs. arXiv e-prints, page arXiv:2003.06079, March 2020.

[20] B. Manuweera, J. Mudge, I. Kahanda, B. Mumey, T. Ramaraj, and A. Cleary. Pangenome-wide association studies with frequented regions. In Proceedings of the 10th ACM International Conference on Bioinformatics, Computational Biology and Health Informatics, pages 627–632. ACM, September 2019.

[21] Mehryar Mohri. Foundations of Machine Learning (Adaptive Computation and Machine Learning). The MIT Press, aug 2012.

[22] C. S. Chin, P. Peluso, et al. Phased diploid genome assembly with single-molecule real-time sequencing. Nat. Methods, 13(12):1050–1054, Dec 2016.

[23] P. D. Sasieni. From genotypes to genes: doubling the sample size. Biometrics, 53(4):1253–1261, Dec 1997.

[24] L.G. Valiant. The complexity of computing the permanent. Theoretical Computer Science, 8(2):189 – 201, 1979.

[25] Lior Rokach and Oded Maimon. Clustering Methods, pages 321–352. Springer US, Boston, MA, 2005.

[26] Ari Kobren, Nicholas Monath, Akshay Krishnamurthy, and Andrew McCallum. A hierarchical algorithm for extreme clustering. In Proceedings of the 23rd ACM SIGKDD International Conference on Knowledge Discovery and Data Mining. ACM, August 2017.

[27] Christos H. Papadimitriou. Computational Complexity. Pearson, ec 1993.

[28] J. Fadista, A. K. Manning, J. C. Florez, and L. Groop. The (in)famous GWAS P-value threshold revisited and updated for low-frequency variants. Eur. J. Hum. Genet., 24(8):1202–1205, 08 2016.

[29] A. M. Molinaro, R. Simon, and R. M. Pfeiffer. Prediction error estimation: a comparison of resampling methods. Bioinformatics, 21(15):3301–3307, Aug 2005.

[30] Chih-Chung Chang and Chih-Jen Lin. LIBSVM: A library for support vector machines. ACM Transactions on Intelligent Systems and Technology, 2:27:1–27:27, 2011.

[31] Eibe Frank, Mark Hall, Len Trigg, Geoffrey Holmes, and Ian H Witten. Data mining in bioinformatics using weka. Bioinformatics, 20(15):2479–2481, October 2004.

[32] A. Auton, L. D. Brooks, et al. A global reference for human genetic variation. Nature, 526(7571):68–74, Oct 2015.

[33] A. Ashley-Koch, Q. Yang, and R. S. Olney. Sickle hemoglobin (HbS) allele and sickle cell disease: a HuGE review. Am. J. Epidemiol., 151(9):839–845, May 2000.

[34] X. Yang, W. P. Lee, K. Ye, and C. Lee. One reference genome is not enough. Genome Biol., 20(1):104, 05 2019.

[35] M. J. Landrum, J. M. Lee, et al. ClinVar: improving access to variant interpretations and supporting evidence. Nucleic Acids Res., 46(D1):D1062–D1067, 01 2018.

[36] P. McColgan and S. J. Tabrizi. Huntington’s disease: a clinical review. Eur. J. Neurol., 25(1):24–34, 01 2018.

[37] J. van de Leemput, J. L. Hess, S. J. Glatt, and M. T. Tsuang. Genetics of Schizophrenia: Historical Insights and Prevailing Evidence. Adv. Genet., 96:99–141, 2016.

[38] M. Balter. Schizophrenia’s Unyielding Mysteries. Sci. Am., 316(5):54–61, Apr 2017.

[39] H. Stefansson, R. A. Ophoff, et al. Common variants conferring risk of schizophrenia. Nature, 460(7256):744–747, Aug 2009.

[40] S. H. Lee, T. R. DeCandia, S. Ripke, J. Yang, P. F. Sullivan, M. E. Goddard, M. C. Keller, P. M. Visscher, and N. R. Wray. Estimating the proportion of variation in susceptibility to schizophrenia captured by common SNPs. Nat. Genet., 44(3):247–250, Feb 2012.

[41] S. M. Purcell, J. L. Moran, et al. A polygenic burden of rare disruptive mutations in schizophrenia. Nature, 506(7487):185–190, Feb 2014.

[42] S. Ripke, B. M. Neale, et al. Biological insights from 108 schizophrenia-associated genetic loci. Nature, 511(7510):421–427, Jul 2014.

[43] Q. Wei and R. L. Dunbrack. The role of balanced training and testing data sets for binary classifiers in bioinformatics. PLoS ONE, 8(7):e67863, 2013.

[44] A. G. Cardno and I. I. Gottesman. Twin studies of schizophrenia: from bow-and-arrow concordances to star wars Mx and functional genomics. Am. J. Med. Genet., 97(1):12–17, 2000.

[45] Y. J. Trakadis, S. Sardaar, A. Chen, V. Fulginiti, and A. Krishnan. Machine learning in schizophrenia genomics, a case-control study using 5,090 exomes. Am. J. Med. Genet. B Neuropsychiatr. Genet., 180(2):103–112, 03 2019.

